# Forecast of omicron wave time evolution

**DOI:** 10.1101/2022.01.16.22269161

**Authors:** R. Schlickeiser, M. Kröger

## Abstract

Adopting an early doubling time of three days for the rate of new infections with the omicron mutant the temporal evolution of the omicron wave in different countries is predicted. The predictions are based on the susceptible-infectious-recovered/removed (SIR) epidemic compartment model with a constant stationary ratio *k* = *μ*(*t*)*/a*(*t*) between the infection (*a*(*t*)) and recovery (*μ*(*t*)) rate. The fixed early doubling time then uniquely relates the initial infection rate *a*_0_ to the ratio *k*, which therefore determines the full temporal evolution of the omicron waves. For each country three scenarios (optimistic, pessimistic, intermediate) are considered and the resulting pandemic parameters are calculated. These include the total number of infected persons, the maximum rate of new infections, the peak time and the maximum 7-day incidence per 100000 persons. Among the considered European countries Denmark has the smallest omicron peak time and the recently observed saturation of the 7-day incidence value at 2478 is in excellent agreement with the prediction in the optimistic scenario. For Germany we predict peak times of the omicron wave ranging from 32 to 38 and 45 days after the start of the omicron wave in the optimistic, intermediate and pessimistic scenario, respectively, with corresponding maximum SDI values of 7090, 13263 and 28911, respectively. Adopting Jan 1st, 2022 as the starting date our predictions implies that the maximum of the omicron wave is reached between Feb 1 and Feb 15, 2022. Rather similar values are predicted for Switzerland. Due to an order of magnitude smaller omicron hospitalization rate, due to the high percentage of vaccinated and boostered population, the German health system can cope with maximum omicron SDI value of 2800 which is about a factor 2.5 smaller than the maximum omicron SDI value 7090 in the optimistic case. By either reducing the duration of intensive care during this period of maximum, and/or by making use of the nonuniform spread of the omicron wave across Germany, it seems that the German health system can barely cope with the omicron wave avoiding triage decisions. The reduced omicron hospitalization rate also causes significantly smaller mortality rates compared to the earlier mutants in Germany. In the optimistic scenario one obtains for the total number of fatalities 7445 and for the maximum death rate 418 per day which are about one order of magnitude smaller than the beta fatality rate and total number.

## I. INTRODUCTION

After being exposed to several Covid-19 outbursts the recently identified omicron mutant threatens many societies wordwide.^1,2^ Not many details are known sofar about its infection characteristics^3,4^ apart from alarming hints (1) that it is spreading at least four times quicker than the beta mutant with a short doubling time of *t*_2_ = 3 days, and (2) that the existing vaccines, taylored to prevent infections from the earlier alpha, beta, gamma and delta mutants, are less efficient against the action of the omicron mutant especially without the current booster campaigns.^5–8^ The alpha (*α*), beta (*β*), gamma (*γ*) and delta (*δ*) mutants have caused the first four Covid-19 waves, respectively. Positively, the omicron mutant seems to lead to on average milder symptons and thus to smaller hospitalization fractions compared to the earlier mutants.

Even with so little details known today it is of high interest to explore quantitatively the future time evolution of the omicron mutant under realistic scenarios of currently taken non-pharmaceutical interventions (NPIs). Of particular interest are reliable estimates of the maximum and total percentage of infected persons from this mutant in order to compare with the available medical capacities in different countries. In the following we provide these estimates by modeling the time evolution of the omicron wave with the susceptible-infectious-recovered/removed (SIR) epidemic compartment model.^9^

As in our earlier analysis^10,11^ – hereafter referred to as KSSIR-model – we adopt a constant stationary ratio *k* = *μ*(*t*)*/a*(*t*) =const. between the infection (*a*(*t*)) and recovery (*μ*(*t*)) rate regulating the transition from susceptible to infected persons and infected to recovered/removed persons in the semi-time case, respectively. As it is so far unclear whether earlier vaccinated persons are not infected by the omicron mutant, we adopt the worst case scenario here and treat the vaccinated persons as fully susceptible to the omicron mutant. However, when calculating hospitalization and mortality rates below we will account for the influence of boosted (with vaccines) persons.

As proven by Table 1 and Figs. 1 and 2 of ref.^10^ the KSSIR-model predicted the temporal evolution of the second wave in several countries convincingly good including the maximum rate 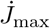 of new infections and the total cumulative number (*J*_∞_) of infections as well as the initial and final second wave time dependence and the time of maximum. For the considered countries the maximum deviation in the total number of infected persons is at most 13 percent off from the later recorded values. An outstanding property of the KSSIR-model is that basically only one parameter, the ratio *k* of recovery and infection rates, fully determines the wave evolution in reduced time *τ*, whereas the influence of the initial fraction *η* of infected people at the onset of the modeled mutant at time *t*_0_ is only minor especially for values of *η* much smaller than unity. Here the reduced time

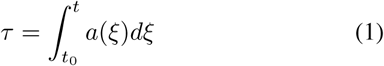

can be calculated for any arbitrary but given real time dependence of the infection rate *a*(*t*).

**TABLE I.**
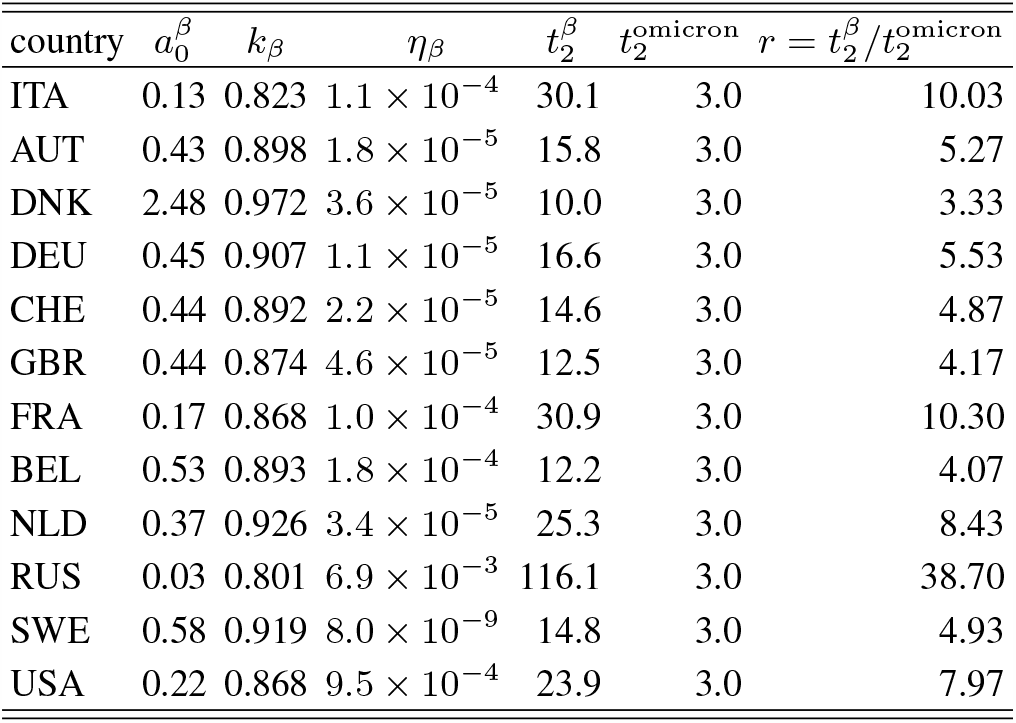
Second wave parameters 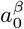 in days^−1^, *k*_*β*_, initial fraction *η*_*β*_, and the inferred second doubling time 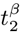 in days. For the omicron mutant in all countries we adopt 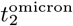 to calculate the ratio of the the two doubling times 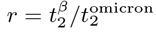.

**FIG. 1.**
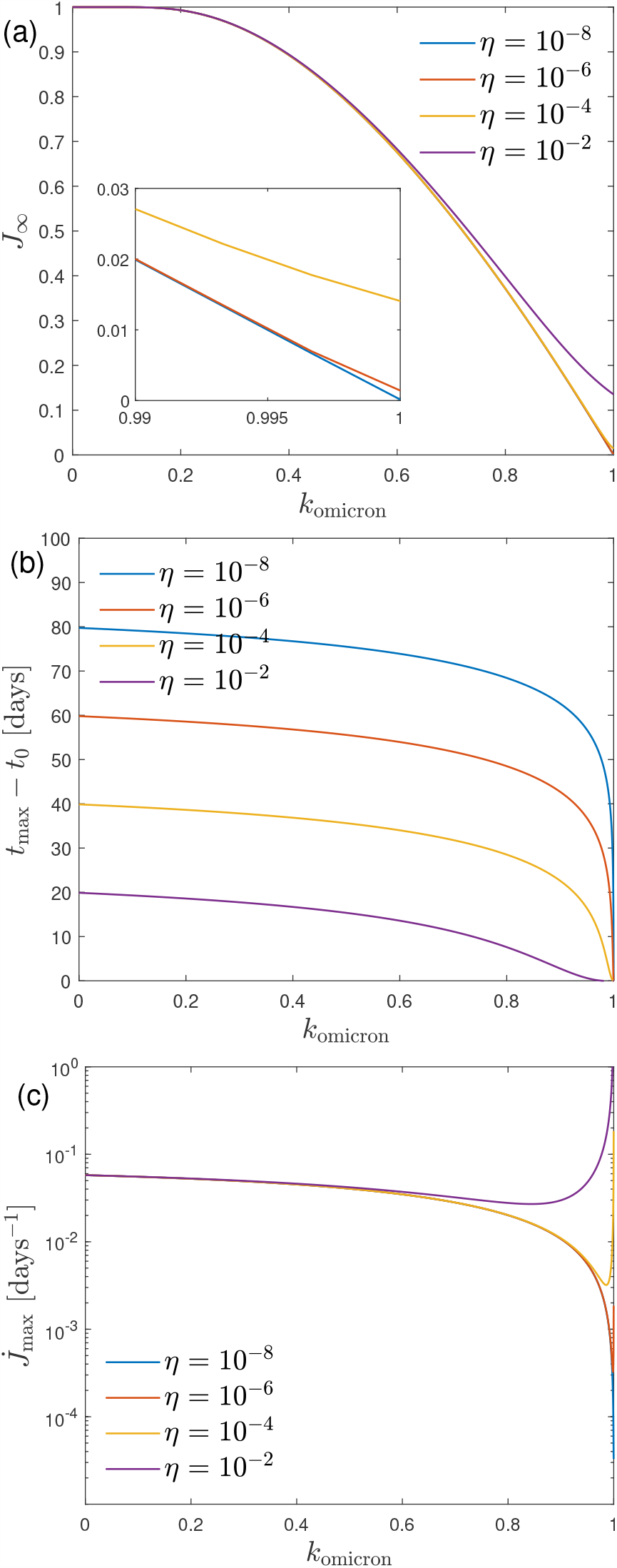
(a) *J*_∞_, (b) *t*_max_ − *t*_0_, and (c) 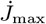 as a function of the only parameter *k* for different values of the initial fraction of infected persons *η* in the case of an early 3-day doubling time.

**FIG. 2.**
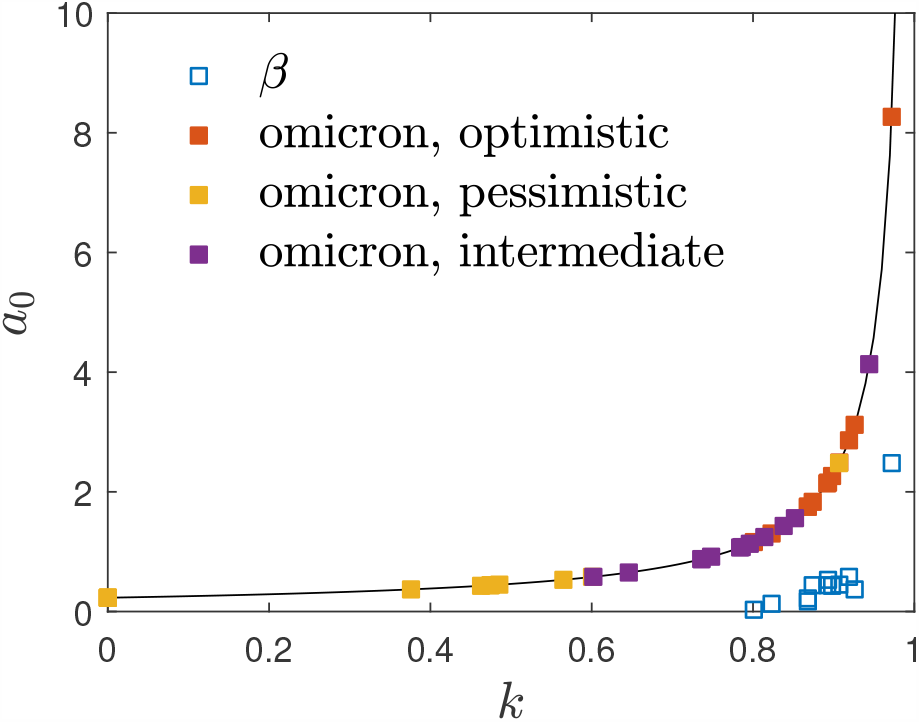
Location of the considered values of *a*_0_ and *k* for the omicron mutant in the investigated 12 countries with the adopted 3-day early doubling times. The symbols *β* represent the values for the earlier beta mutant. The solid black line is Eq. (23).

Adopting a constant infection rate *a*(*t*) = *a*_0_ is a good approximation for rapidly evolving mutant waves and not only for its initial phases, so that in this case the simple relation *τ* = *a*_0_(*t* − *t*_0_) holds between the reduced and the real time. Moreover, by determining the then two decisive parameters *k* and *a*_0_ from the early monitored real time evolution then allows us the accurate determination of all relevant quantities of the considered outburst. The two parameters *k* and *a*_0_ differ among different societies depending besides specific virus mutant properties also on the NPIs taken, the quality and ability of the health care system, and the discipline of the people in keeping distances, wearing masks and following quarantine measures. As the latter are mainly unchanged during different mutant actions it makes sense to relate the *k* and *a*_0_ parameters of the omicron mutant to those of the earlier beta mutant as we will adopt below.

## II. RESULTS FROM THE SIR-MODEL

In terms of the reduced time (1) the KSSIR model equations read

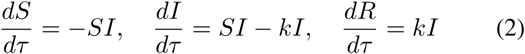

obeying the sum constraint

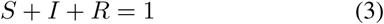

at all times. In Eqs. (2)-(3) *S, I* and *R* denote the fractions of susceptible, infected and recovered/removed persons in a population, respectively, subject to the semi-time initial conditions

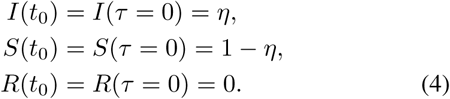

The rate of new infections and its cotresponding cumulative number are given by *j*(*τ*) = *S*(*τ*)*I*(*τ*) and 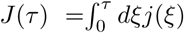, respectively, whereas 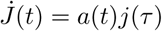 and *J*(*t*) = *J*(*τ*).

### A. Exact results

In terms of *J* the exact solution of the KSSIR model in the semi-time case is given by^11^

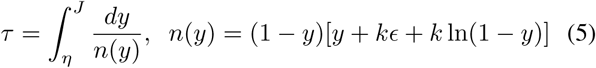

with *ϵ* = − ln(1 − *η*). The remaining SIR quantities are given by *J*(*τ*) as *S*(*τ*) = 1 − *J*(*τ*), *I*(*τ*) = *J*(*τ*)+*kϵ*+*k* ln[1 − *J*(*τ*)] and *R*(*τ*) = − *k*[*ϵ* + ln(1 − *J*(*τ*))]. Differentiating Eq. (5) with respect to *τ* readily yields for the rate of new cases

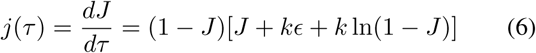

As shown before^11^ without the explicit inversion of the solution (5) to *J*(*τ*) one obtains for the final cumulative fraction of infected persons

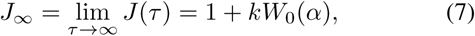

with *α* = − (1 − *η*)*k*^−1^*e*^−1*/k*^, and for the maximum rate of new infections

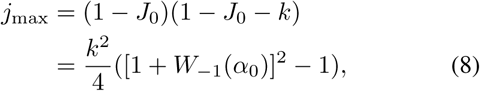

occurring at

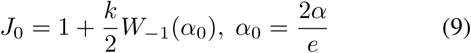

in terms of the principal (*W*_0_) and non-principal (*W*_−1_) solution of Lambert’s equation^12^, the well-known and documented Lambert functions. We emphasize that for small values of *η* ≪ 1 the results (7) and (9) are basically independent of the value of *η* and only determined by the parameter *k*. The first Eq. (8) implies

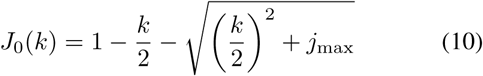

### B. Approximate results

Very accurate approximations have been obtained^12^ for

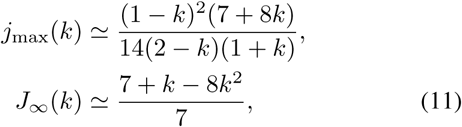

so that Eq. (10) provides the approximation

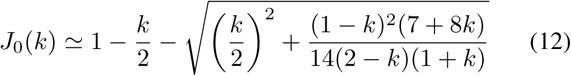

Note that for small *k* < 1*/*8 the exact expressions (7), (9), and (10) are still useful as the approximation gives values slightly larger than unity for *J*_∞_.

The occurrence of the maximum rate of new infections (8) at positive values of the reduced peak time *τ*_max_ > 0 requires values of *k* < 1 −2*η*. In this case^11^ the reduced peak time is well approximated by

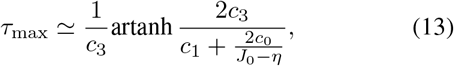

with *c*_0_ = *η*(1 − *η*) and *c*_1_ = 1 − *k* − 2*η*,

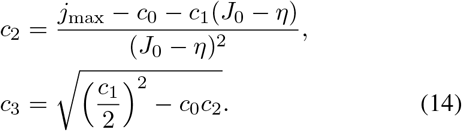

The reduced time dependence of the rate of new infections is well approximated as

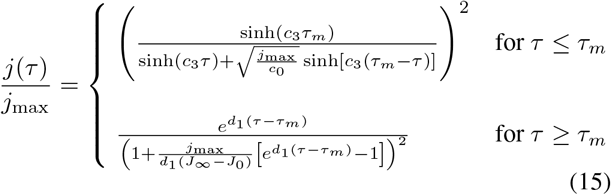

with *d*_1_ = *J* _∞_ − (1 − *k*).

For a stationary infection rate *a*_0_ the corresponding real peak time is given by

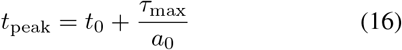

Likewise, the early asymptotic reduced time behavior is well approximated^12^ by

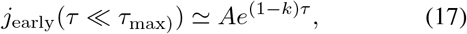

corresponding to the early asymptotic real time behavior

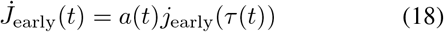

In the considered case of a stationary infection rate Eq. (18) reduces to

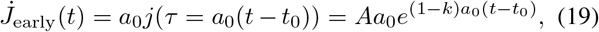

implying for the early doubling time defined by 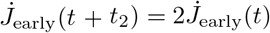 that

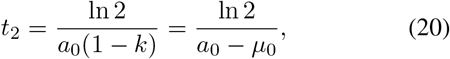

where we inserted *k* = *μ*_0_*/a*_0_ in the case of stationary infection and recovery rates. Equation (20) will be used in the following two sections in two different ways.

The maximum 7-day incidence value per 10^5^ persons is calculated by integrating

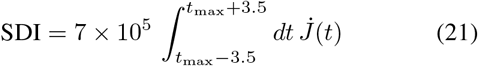

It is only slightly smaller than the estimate SDI ≃ 7 × 10^5^ 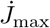 from the maximum rate.

The late at times after the maximum half-decay time is given by

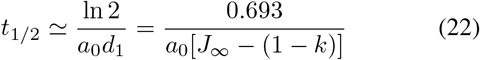

## III. CONSEQUENCES OF EARLY 2-DAY DOUBLING TIME

For the omicron mutant the early doubling time of *t*_2,omicron_ = 3 days has been reported^3,4^ in South Africa, Great Britain and Denmark. Adopting this value for all countries considered then provides according to Eq. (20) for the omicron mutant the relation

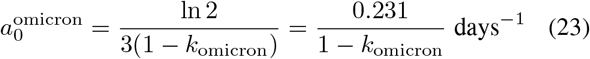

throughout. Using this relation in all results of the last section to eliminate *a*_0_ we find that all quantities of interest are solely determined by the parameter *k*. Particularly for the peak time (16) we obtain

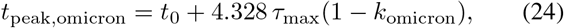

whereas the maximum rate of new infections

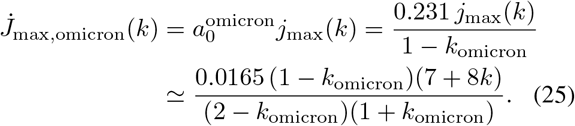

Likewise the real time dependence of the rate of new infections with Eq. (14) is given by

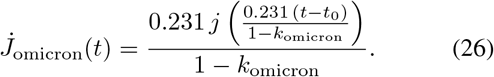

In Fig. 1 we display the resulting dependence of *J*_∞_, 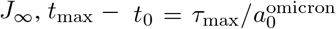 and 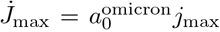 as a function of the parameter *k*_omicron_ ∈ [0, 1]. It can be seen that *J*_∞_ and 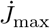 decrease with increasing values of *k* almost independent of the initial fraction *η* of infected persons, except at very large values of *k*_omicron_ close to unity. Obviously, for comparatively small values of the total number of infected persons *J*_∞_ and the maximum rate of newly infected persons 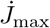 large values of the ratio *k* are required. Alternatively, the reduced time of maximum *τ*_max_ decreases with increasing values of *k* as long as *k* is much smaller than 1 − 2*η*.

## IV. OMICRON FORECAST IN INDIVIDUAL COUNTRIES

With the earlier inferred parameter values^10^ 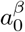 and *k*_*β*_ for the second wave caused by the *β*-mutant we calculate the second wave doubling time 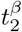 for the countries listed in Table I. Adopting for all countries the short omicron doubling time 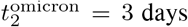 we obtain the ratio of the doubling times also listed in the last column of Table I. With Eq. (20) we then infer for this ratio

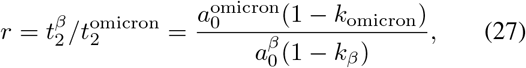

yielding readily the relation

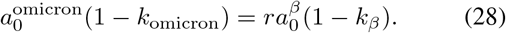

For each country we then consider 3 possible omicron scenarios:

1. the *optimistic* case with *k*_omicron_ = *k*_*β*_ so that the increase in the ratio *r* is solely due to the increase in the stationary infection rate

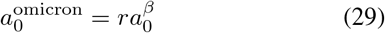 As noted earlier the larger the value of *k*_omicron_ the smaller the total cumulative number of infections *J*_∞_ and the maximum rate of new infections *j*_max_ will be. This justifies the classification of this case as optimistic.
2. the *pessimistic* scenario with 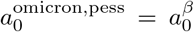 so that the increase in the ratio *r* is solely due to the decrease in the ratio *k*

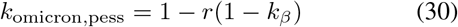 Clearly, with these small values of *k*_omicron_ the resulting total cumulative number of infections *J*_∞_ and the maximum rate of new infections *j*_max_ will be highest, justifying the classification of this case as pessimistic. In four countries (ITA, FRA, RUS, USA) the resulting *k*_omicron,pess_ is negative which cannot be. In these cases we use *k*_omicron,pess_ = 0 and 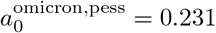.
3. the *intermediate* case with

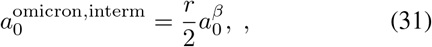

where half of the increase in the ratio *r* stems from the increase in the stationary infection rate. Then as a consequence

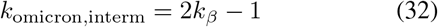

In Tables II, III and IV we calculate the forecast for the omicron mutant for these three scenarios, respectively. Figure 2 visualizes the relationship between *a*_0_ and *k* and the location of the three regimes for the 12 countries, and Fig. 3 shows the time dependence of 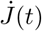 and cumulative fraction *J*(*t*) of infected persons for all 12 countries.

**TABLE II.** Forecast of the omicron mutant for the optimistic case, i.e., 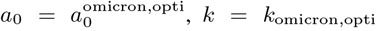, and initial fraction *η* = *η*_*β*_ from Tab. I for this table. Columns list the final cumulative fraction *J*_∞_ of infected persons, the maximum (dimensionless) rate *j*_max_ of new infections, the cumulative fraction *J*_0_ of infected persons at peak time, the reduced peak time *τ*_max_, the peak time *t*_max_ − *t*_0_ in days, and the SDI, the maximum 7-day incidence per 10^5^ persons. Country names are abbreviated by their *α*_3_ codes.

| optimistic scenario |  |  |  |  |  |  |  |  |  |  |
| --- | --- | --- | --- | --- | --- | --- | --- | --- | --- | --- |
| $\alpha_3$ | $a_0$ | $k$ | $J_\infty$ | $j_{\text{max}}$ | $\dot{J}_{\text{max}}$ | $J_0$ | $\tau_{\text{max}}$ | $t_{\text{max}} - t_0$ | SDI | |
| ITA | 1.304 | 0.823 | 0.33 | 0.0139 | 0.018 | 0.160 | 35.6 | 27 days | 12725 |  |
| AUT | 2.265 | 0.898 | 0.20 | 0.0049 | 0.011 | 0.097 | 68.9 | 30 days | 7716 |  |
| DNK | 8.267 | 0.972 | 0.06 | 0.0004 | 0.004 | 0.028 | 130.1 | 16 days | 2424 |  |
| DEU | 2.490 | 0.907 | 0.18 | 0.0041 | 0.010 | 0.089 | 79.0 | 32 days | 7090 |  |
| CHE | 2.141 | 0.892 | 0.21 | 0.0054 | 0.012 | 0.102 | 64.0 | 30 days | 8148 |  |
| GBR | 1.833 | 0.874 | 0.24 | 0.0073 | 0.013 | 0.118 | 51.3 | 28 days | 9392 |  |
| FRA | 1.751 | 0.868 | 0.25 | 0.0080 | 0.014 | 0.123 | 43.5 | 25 days | 9853 |  |
| BEL | 2.155 | 0.893 | 0.21 | 0.0055 | 0.012 | 0.101 | 44.3 | 21 days | 8250 |  |
| NLD | 3.120 | 0.926 | 0.14 | 0.0026 | 0.008 | 0.071 | 77.4 | 25 days | 5751 |  |
| RUS | 1.161 | 0.801 | 0.39 | 0.0219 | 0.025 | 0.173 | 10.7 | 9 days | 17773 |  |
| SWE | 2.861 | 0.919 | 0.16 | 0.0031 | 0.009 | 0.078 | 175.6 | 61 days | 6216 |  |
| USA | 1.753 | 0.868 | 0.26 | 0.0087 | 0.015 | 0.122 | 26.0 | 15 days | 10650 |  |

**TABLE III.**
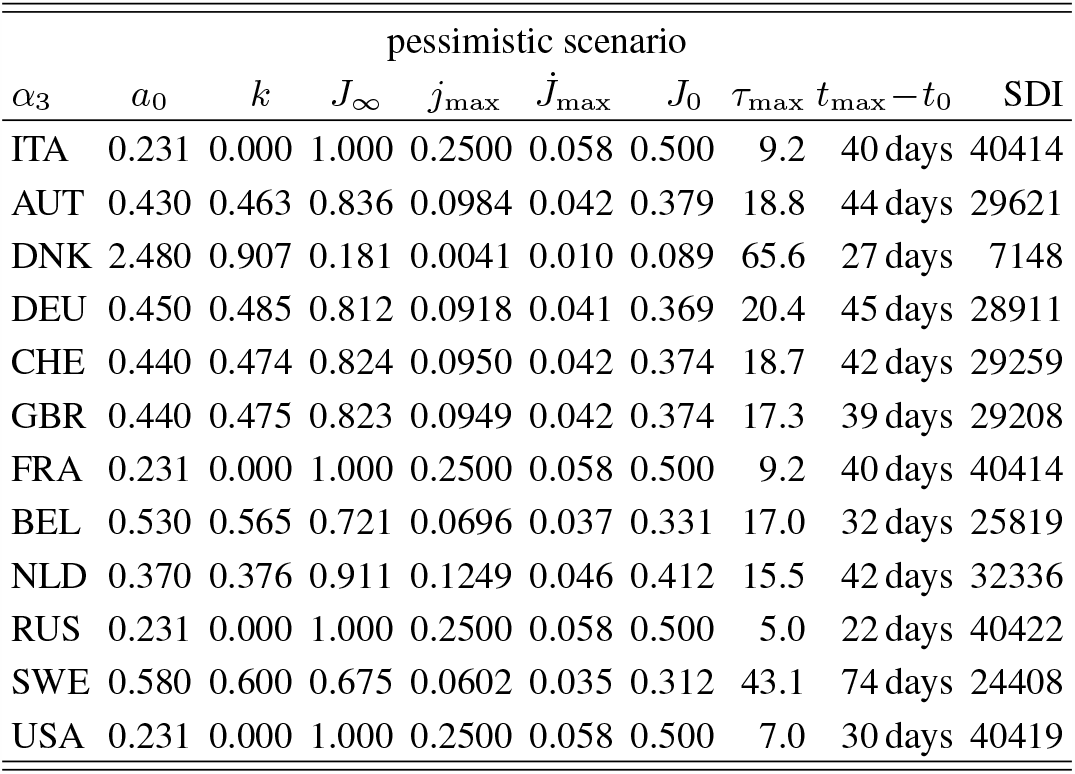
Forecast of the omicron mutant for the pessimistic case, i.e., 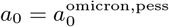 and *k* = *k*_omicron,pess_.

| pessimistic scenario |  |  |  |  |  |  |  |  |  |  |
| --- | --- | --- | --- | --- | --- | --- | --- | --- | --- | --- |
| $\alpha_3$ | $a_0$ | $k$ | $J_\infty$ | $j_{\text{max}}$ | $\dot{J}_{\text{max}}$ | $J_0$ | $\tau_{\text{max}}$ | $t_{\text{max}} - t_0$ | SDI | |
| ITA | 0.231 | 0.000 | 1.000 | 0.2500 | 0.058 | 0.500 | 9.2 | 40 days | 40414 |  |
| AUT | 0.430 | 0.463 | 0.836 | 0.0984 | 0.042 | 0.379 | 18.8 | 44 days | 29621 |  |
| DNK | 2.480 | 0.907 | 0.181 | 0.0041 | 0.010 | 0.089 | 65.6 | 27 days | 7148 |  |
| DEU | 0.450 | 0.485 | 0.812 | 0.0918 | 0.041 | 0.369 | 20.4 | 45 days | 28911 |  |
| CHE | 0.440 | 0.474 | 0.824 | 0.0950 | 0.042 | 0.374 | 18.7 | 42 days | 29259 |  |
| GBR | 0.440 | 0.475 | 0.823 | 0.0949 | 0.042 | 0.374 | 17.3 | 39 days | 29208 |  |
| FRA | 0.231 | 0.000 | 1.000 | 0.2500 | 0.058 | 0.500 | 9.2 | 40 days | 40414 |  |
| BEL | 0.530 | 0.565 | 0.721 | 0.0696 | 0.037 | 0.331 | 17.0 | 32 days | 25819 |  |
| NLD | 0.370 | 0.376 | 0.911 | 0.1249 | 0.046 | 0.412 | 15.5 | 42 days | 32336 |  |
| RUS | 0.231 | 0.000 | 1.000 | 0.2500 | 0.058 | 0.500 | 5.0 | 22 days | 40422 |  |
| SWE | 0.580 | 0.600 | 0.675 | 0.0602 | 0.035 | 0.312 | 43.1 | 74 days | 24408 |  |
| USA | 0.231 | 0.000 | 1.000 | 0.2500 | 0.058 | 0.500 | 7.0 | 30 days | 40419 |  |

**TABLE IV.** Forecast of the omicron mutant for the intermediate case, i.e., 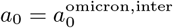 and *k* = *k*_omicron,inter_.

| intermediate scenario |  |  |  |  |  |  |  |  |  |  |
| --- | --- | --- | --- | --- | --- | --- | --- | --- | --- | --- |
| $\alpha_3$ | $a_0$ | $k$ | $J_\infty$ | $j_{\text{max}}$ | $\dot{J}_{\text{max}}$ | $J_0$ | $\tau_{\text{max}}$ | $t_{\text{max}} - t_0$ | SDI | |
| ITA | 0.652 | 0.646 | 0.613 | 0.0489 | 0.032 | 0.286 | 21.4 | 33 days | 22307 |  |
| AUT | 1.132 | 0.796 | 0.378 | 0.0181 | 0.021 | 0.182 | 41.0 | 36 days | 14328 |  |
| DNK | 4.133 | 0.944 | 0.110 | 0.0015 | 0.006 | 0.054 | 90.9 | 22 days | 4462 |  |
| DEU | 1.245 | 0.814 | 0.347 | 0.0152 | 0.019 | 0.168 | 46.7 | 38 days | 13263 |  |
| CHE | 1.071 | 0.784 | 0.398 | 0.0201 | 0.022 | 0.191 | 38.2 | 36 days | 15060 |  |
| GBR | 0.917 | 0.748 | 0.458 | 0.0267 | 0.024 | 0.218 | 31.0 | 34 days | 17109 |  |
| FRA | 0.876 | 0.736 | 0.477 | 0.0290 | 0.025 | 0.226 | 26.8 | 31 days | 17797 |  |
| BEL | 1.078 | 0.786 | 0.396 | 0.0199 | 0.021 | 0.189 | 28.6 | 27 days | 14979 |  |
| NLD | 1.560 | 0.852 | 0.281 | 0.0099 | 0.016 | 0.137 | 48.0 | 31 days | 10832 |  |
| RUS | 0.580 | 0.602 | 0.678 | 0.0627 | 0.036 | 0.307 | 8.8 | 15 days | 25466 |  |
| SWE | 1.431 | 0.838 | 0.305 | 0.0117 | 0.017 | 0.148 | 96.2 | 67 days | 11748 |  |
| USA | 0.876 | 0.736 | 0.479 | 0.0295 | 0.026 | 0.226 | 18.3 | 21 days | 18110 |  |

**FIG. 3.**
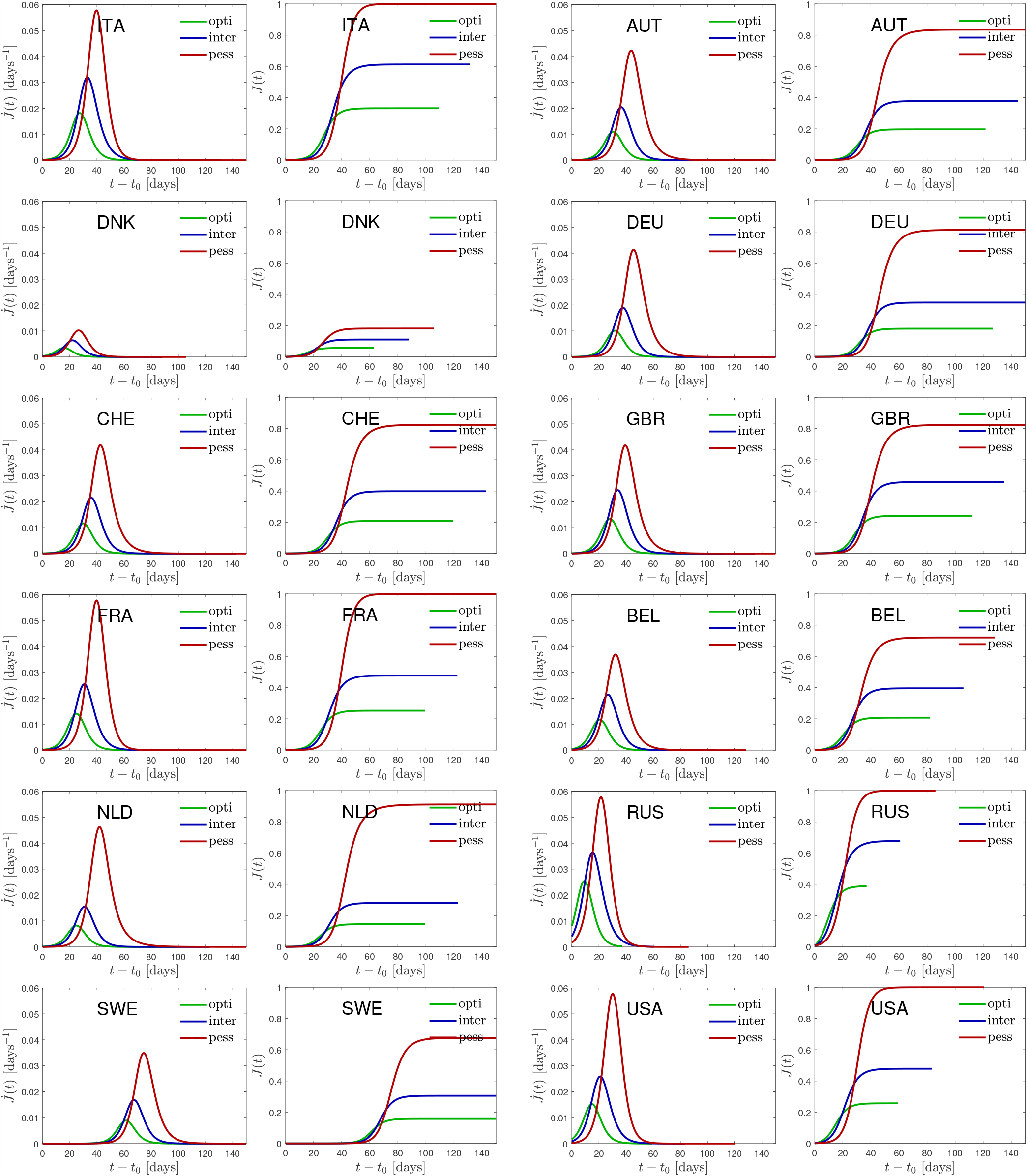
Time dependence of the daily rate of newly infected persons, 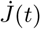, as well as the cumulative fraction of infected persons, *J* (*t*), for all 12 countries.

It is obvious from these three tables that in European countries, apart from Russia with limited data reliability, Denmark has the shortest peak time of the omicron wave ranging from 16 to 22 and 27 days after the start of the omicron wave in the optimistic, intermediate and pessimistic scenario, respectively. The corresponding predicted maximum 7-day incidence values per 10^5^ persons (SDI) are 2424, 4462 and 7148, respectively. Presently on January 10, 2022 the well-monitored data of Denmark^13^ indicate that the SDI has saturated at its maximum value at 2478 which is in excellent agreement with our predicted value in the optimistic case. Although preliminary this outstanding agreement is definitely encouraging and argues in favour of the optimistic scenario

Regarding Germany we predict peak times of the omicron wave ranging from 32 to 38 and 45 days after the start of the omicron wave in the optimistic, intermediate and pessimistic scenario, respectively, with corresponding maximum SDI values of 7090, 13263 and 28911, respectively. Adopting Jan 1st, 2022 as the starting date our predictions implies that the maximum of the omicron wave is reached between Feb 1 and Feb 15, 2022. In the optimistic case the total cumulative number of omicron infections will be 0.180 but can go up high to 0.812 in the pessimistic case. The late half decay times are 3.2 to 3.5 and 5.2 days in the optimistic, intermediate and pessimistic case, respectively.

Rather similar values are predicted for Switzerland. Here the peak times of the omicron wave range from 30 to 36 and 42 days after the start and the corresponding maximum SDI values are 8148, 15060 and 29259, respectively. Here with the same starting date the maximum of the omicron wave is reached between Jan 31 and Feb 13, 2022. Here, in the optimistic case the total cumulative number of omicron infections will be 0.208 but can go up high to 0.824 in the pessimistic case. The late half decay times are 3.2 to 3.6 and 5.3 days in the optimistic, intermediate and pessimistic case, respectively.

## V. MEDICAL CONSEQUENCES FOR GERMANY

### A. Tolerable maximum 7-day incidence value

We have argued earlier^14^ that the German health system can cope with maximum SDI values of 280*/*(*hm*) without any triage decisions, where *m* in months denotes the average duration of intensive care with access to breathing apparati for seriously infected persons, and *h* in units of percent indicates the percentage of people seriously infected needing access to breathing apparati in hospitals. For the earlier *α* and *β* mutants the value of *h* = 1 has been reasonable. Fortunately, for the omicron variant from studies in South Africa^15^ and Great Britain^16^ substantial 70–90 percent reductions as compared to earlier mutants in Covid-19 hospitalization have been reported. This strong reduction is predominantly caused by the high percentage of persons with boostered vaccination.

We therefore adopt here the value of *h*_omicron_ = 0.1, i.e. only one out of 1000 new infections with the omicron mutant needs to be hospitalised. Consequently, the German health system can cope with maximum omicron SDI value of 2800*/m* which is about a factor 2.5 smaller than the maximum omicron SDI value 7090 in the optimistic case. By either (1) reducing the duration of intensive care during this period of maximum to *m* = 0.5, and/or (2) by making use of the nonuniform spread of the omicron wave across Germany, appearing first in the northern states and considerably later in the southern and eastern states, combined with mutual help in hospital capacities, it seems that the German health system can cope with the omicron wave avoiding triage decisions.

### B. Fatality rates and total number of fatalities

As before^10,14^ we assume that every second hospitalised person eventually dies from the omicron virus so that the omicron mortality rate is *f*_omicron_ = 0.5*h*_omicron_ = 5 × 10^−4^ which is one order of magnitude smaller than the mortality rates of the earlier mutants. Consequently, the total fatality rate is given by *D*_∞_ = *f*_omicron_*J*_∞_*N*, where *N* = 82.7 million denotes the German population. Likewise the maximum death rate is 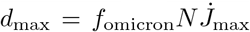. In the optimistic scenario one obtains *D*_∞_ = 7445 and *d*_max_ = 418 per day which are about one order of magnitude smaller than the beta fatality rate and total number of fatalities of the second wave^10^. The main reason for these comparatively small numbers is the order of magnitude smaller hospitalization rate of the omicron mutant compared to the earlier more deadly mutants.

However, in the less likely pessimistic scenario the fatality numbers increase by a factor 4.5 to to *D*_∞_ = 33576 and *d*_max_ = 1708 which are about half of the fatality values of the second wave.

## VI. SUMMARY AND CONCLUSIONS

Adopting an early doubling time of three days for the rate of new infections with the omicron mutant the temporal evolution of the omicron wave in different countries is predicted. The predictions are based on the susceptible-infectious-recovered/removed (SIR) epidemic compartment model with a constant stationary ratio *k* = *μ*(*t*)*/a*(*t*) between the infection (*a*(*t*)) and recovery (*μ*(*t*)) rate. The fixed early doubling time then uniquely relates the initial infection rate *a*_0_ to the ratio *k*, which therefore determines the full temporal evolution of the omicron waves.

As all considered countries have been exposed to earlier waves of the Covid-19 virus we relate the parameters *a*_0_ and *k* to those of the well-studied second wave. In the optimistic case we assume that the decrease in the early doubling time of the omicron mutant as compared to the beta mutant is solely due to a corresponding increase in the initial infection rate *a*_0_ whereas the ratio *k* is the same as for the beta mutant. In the pessimistic case we assume that the decrease in the early doubling times is fully caused by a corresponding decrease in the ratio *k* whereas the initial infection rate is the same as for the beta mutant. In the intermediate scenario half of the decrease in the early doubling time is assigned to a corresponding increase of the initial infection rate and a corresponding decrease of the ratio *k*. For 12 countries these three scenarios (optimistic, pessimistic, intermediate) are considered and the resulting pandemic parameters are calculated. These include the total number of infected persons, the maximum rate of new infections, the peak time and the maximum 7-day incidence per 100000 persons.

Among the considered European countries Denmark has the smallest omicron peak time and the recently observed saturation of the 7-day incidence value at 2478 is in excellent agreement with our prediction in the optimistic case. For Germany we predict peak times of the omicron wave ranging from 32 to 38 and 45 days after the start of the omicron wave in the optimistic, intermediate and pessimistic scenario, respectively, with corresponding maximum SDI values of 7090, 13263 and 28911, respectively. Adopting Jan 1st, 2022 as the starting date our predictions implies that the maximum of the omicron wave is reached between Feb 1 and Feb 15, 2022. In the optimistic case the total cumulative number of omicron infections will be 0.180 but can go up high to 0.812 in the pessimistic case. The late half decay times are 3.2 to 3.5 and 5.2 days in the optimistic, intermediate and pessimistic case, respectively.

Rather similar values are predicted for Switzerland. Here the peak times of the omicron wave range from 30 to 36 and 42 days after the start and the corresponding maximum SDI values are 8148, 15060 and 29259, respectively. Here with the same starting date the maximum of the omicron wave is reached between Jan 31 and Feb 13, 2022. Here, in the optimistic case the total cumulative number of omicron infections will be 0.208 but can go up high to 0.824 in the pessimistic case. The late half decay times are 3.2 to 3.6 and 5.3 days in the optimistic, intermediate and pessimistic case, respectively.

Adopting an order of magnitude smaller omicron hospitalization rate thanks to the high percentage of vaccinated and boostered population we conclude that the German health system can cope with maximum omicron SDI value of 2800 which is about a factor 2.5 smaller than the maximum omicron SDI value 7090 in the optimistic case. By either reducing the duration of intensive care during this period of maximum, and/or by making use of the nonuniform spread of the omicron wave across Germany, it seems that the German health system can barely cope with the omicron wave avoiding triage decisions.

The reduced omicron hospitalization rate also causes significantly smaller mortality rates compared to the earlier mutants in Germany. In the optimistic scenario one obtains for the total number of fatalities *D*_∞_ = 7445 and for the maximum death rate *d*_max_ = 418 per day which are about one order of magnitude smaller than the beta fatality rate and total number. In the less likely pessimistic scenario these numbers increase by a factor 4.5.

## Data Availability

All data produced in the present work are contained in the manuscript

## References

1. K. Kupferschmidt and G. Vogel, Covid-19 how bad is omicron? some clues are emerging, Science 374, 1304 (2021).

2. T. Nature Editors, The global response to omicron is making things worse, Nature 600, 190 (2021).

3. R. Viana, S. Moyo, D. G. Amoako, and et al., Rapid epidemic expansion of the sars-cov-2 omicron variant in southern africa, medRxiv, medrxiv (2021).

4. R. C. Barnard, N. G. Davies, C. A. B. Pearson, M. Jit, and W. J. Edmunds, Projected epidemiological consequences of the omicron sars-cov-2 variant in england, december 2021 to april 2022, medRxiv 10.1101/2021.12.15.21267858 (2021).

5. C. C. R. T. Perrine, C. G., Sars-cov-2 b.1.1.529 (omicron) variant - united states, december 1-8, 2021, MMWR-Morbidity and Mortality weekly report 70, 1731 (2021).

6. S. S. A. Karim and Q. A. Karim, Omicron sars-cov-2 variant: a new chapter in the covid-19 pandemic, Lancet 398, 2126 (2021).

7. E. Mahase, Covid-19: Do vaccines work against omicron-and other questions answered, BMJ-Brit. Med. J. 375, n3062 (2021).

8. I. Torjesen, Covid restrictions tighten as omicron cases double every two to three days, BMJ-Brit. Med. J. 375, n3051 (2021).

9. E. Estrada, Covid-19 and sars-cov-2. modeling the present, looking at the future, Phys. Rep. 869, 1 (2020).

10. M. Kröger and R. Schlickeiser, Verification of the accuracy of the SIR model in forecasting based on the improved sir model with a constant ratio of recovery to infection rate by comparing with monitored second wave data, R. Soc. Open Sci. 8, 211379 (2021).

11. R. Schlickeiser and M. Kröger, Analytical solution of the SIR-model for the temporal evolution of epidemics: part b. semi-time case, J. Phys. A 54, 175601 (2021).

12. M. Kröger and R. Schlickeiser, Analytical solution of the SIR-model for the temporal evolution of epidemics. part a: Time-independent reproduction factor, J. Phys. A 53, 505601 (2020).

13. Editors, Worldometers.info (Dover, Delaware, USA, 2022) accessed on 11 Jan 2022, https://www.worldometers.info/coronavirus/country/denmark/.

14. R. Schlickeiser and M. Kröger, Reasonable limiting of 7-day incidence per hundred thousand and herd immunization in germany and other countries, Covid 1, 130 (2021).

15. N. Wolter, W. Jassat, and S. Walaza, Early assessment of the clinical severity of the sars-cov-2 omicron variant in south africa, medRxiv 10.1101/2021.12.21.21268116 (2021).

16. A. Sheikh, S. Kerr, M. Woolhouse, J. McMenamin, and C. Robertson, Severity of omicron variant of concern and vaccine effectiveness against symptomatic disease: national cohort with nested test negative design study in scotland, The University of Edinburgh preprint server 22 Dec (2021), https://www.research.ed.ac.uk/en/publications.

